# Fusion of middle ear optical coherence tomography and computed tomography in three ears

**DOI:** 10.1101/2024.09.23.24314125

**Authors:** Junzhe Wang, Floor Couvreur, Joshua D. Farrell, Reshma Ghedia, Nael Shoman, David P Morris, Robert B.A. Adamson

**Affiliations:** School of Biomedical Engineering, Dalhousie University, NS B3H 4R2, Canada; Division of Otolaryngology-Head and Neck Surgery, Department of Surgery, Dalhousie University, Halifax, NS B3H 4R2, Canada; Department of Otorhinolaryngology, Head and Neck Surgery, AZ Sint-Jan’s Hospital, Ruddershove 10, 8000 Bruges, Belgium; Electrical and Computer Engineering Department, Dalhousie University, Halifax, NS B3H 4R2, Canada

## Abstract

**Importance:** Middle ear OCT imaging in patients has not previously been directly compared to a standard of care clinical 3D imaging technology such as CT. This represents the first such comparison and provides new insight into OCT’s capabilities, strengths and limitations.

**Objective:** To qualitatively compare the capabilities of middle ear OCT to CT in normal and pathological ears on representative slices in co-registered OCT and CT datasets.

**Design, Setting, and Participants:** One normal middle ear, one ear affected by traumatic injury and one ear with cholesteatoma were imaged with both OCT and high-resolution clinical temporal bone CT. Participants were drawn from the patient population of a tertiary otology clinic. CT and OCT images were aligned using rigid co-registration with manual landmark selection.

**Main Outcomes and Measures:** Images were analyzed qualitatively for field of view, resolution, shadowing, artefacts, soft tissue and bony tissue contrast and presentation of diagnostically important features.

**Results:** In the three imaged ears, OCT was capable of visualizing many of the important features indicative of middle ear pathology. When compared to CT, OCT was found to exhibit a limited field of view (FOV) largely confined to the mesotympanum and subject to shadowing from bony structures. However, OCT could resolve soft tissue features that were not readily apparent in the CT images, to have a higher resolution than CT and to provide excellent anatomical fidelity with CT which allowed OCT images to be accurately co-registered with CT images.

**Conclusions and Relevance:** The results support a role for middle ear OCT in otological diagnostics. While OCT is not capable of replacing CT due to its limited FOV and inability to image through thick bony tissues, it can visualize many signs of pathology including some soft tissue features that are difficult to visualize with CT. Given OCT’s ability to image in real-time, its compatibility with in-office imaging and its lack of ionizing radiation, it may, despite its limitations compared to CT, be an appealing imaging modality for many applications in middle ear diagnostics.

**Key Points:** *Question (1 sentence):* What are the strengths and limitations of middle ear optical coherence tomography (OCT) imaging compared to computed tomography (CT) in real-world clinical imaging scenarios?

*Finding (1-2 sentence):* OCT and CT imaging produce complementary diagnostic information with CT offering unobstructed images of bony anatomy and OCT providing the ability to visualize soft tissue.

*Meaning (1 sentence):* OCT, CT and fused OCT/CT imaging can each provide useful, complementary diagnostic information in clinical otology.

## Introduction

High resolution computed tomography (CT) imaging is a mainstay of modern middle ear diagnostics, long serving as the preferred non-invasive visualization modality for diagnosing ossicular pathologies and for surgical planning^1–3^. Middle ear optical coherence tomography (ME-OCT) is an emerging^4^, point-of-care diagnostic technology capable of providing volumetric images of the middle ear space through the intact tympanic membrane (TM) without exposing the patient to ionizing radiation^5–11^. These two imaging modalities are complementary in several respects^9,11^. CT enables visualization of osseous structures^2,3,12^ anywhere in the middle ear without shadowing artefacts^4,7^. However, CT is not real-time, is ionizing, has difficulty resolving fine soft tissue structures and provides lower resolution than OCT imaging^1,13–15^. ME-OCT provides radiation-free, video-rate real-time imaging and can resolve fine soft tissue structures like the TM^9,16^. However, being a line-of-sight modality^4,6,9^, ME-OCT imaging can be obstructed by bone or soft tissue thicker than approximately 1mm and so it is limited to the field of view (FOV) visible through the TM^4,7,17^. ME-OCT also exhibits artefacts from shadowing and multiple scattering that do not affect CT images^4,7,11,18^. Both modalities are capable of producing images with high geometric fidelity to the patient anatomy and so CT and OCT images of the same anatomy can be co-registered through rigid transformations^9,19,20^.

In this study we provide, for the first time, a superimposed comparison of volumetric images taken with OCT and clinical CT in the same ears. Images are presented for three patient ears: a normal middle ear, an ear affected by a traumatic injury and a chronic ear with a cholesteatoma. In each case, we examine the strengths and limitations of the two modalities and discuss how they can complement each other to provide new diagnostic insight when combined.

OCT is an optical interferometric imaging modality in which light reflected from a patient ear is interfered with reference light and the intensity of the interfered light is analyzed^9,21–23^ to produce a depth-resolved reflectivity profile of middle ear structures along the line of sight ^16,18^. By scanning the sample beam along a line or over a surface, image lines can be assembled into 2D and 3D images of tissue with an appearance similar to an ultrasound image ^4,24,25^. We have developed and previously described a turnkey, handheld, real-time ME-OCT research system (Figure 1A) comprising an otoscopic handpiece tethered to a console containing a swept-source laser, interferometer and image-processing software^9,16,26–28^. The handpiece contains a video otoscope, a ME-OCT scanner and a speaker for presenting tone stimuli for OCT Doppler vibrometry (a capability not used in the current study but described elsewhere^16,26,29^). The system operates at 1550nm and achieves an axial resolution of 37.5μm, a lateral resolution of 50μm and an imaging depth of 13mm^9^.

**Figure 1.**
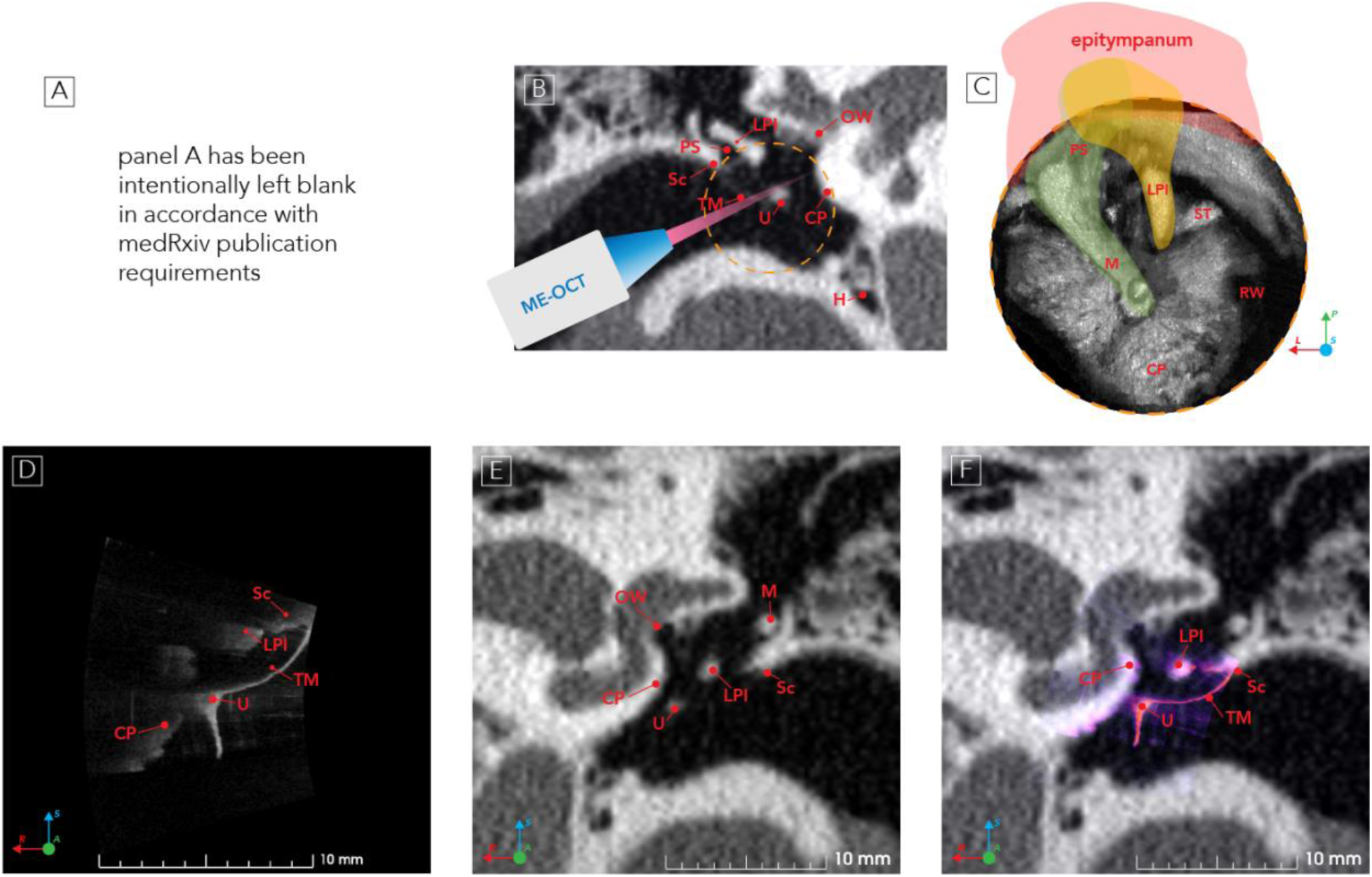
Middle Ear OCT Acquisition and Fusion with CT. A, Image of a clinician using the OCT handpiece to image the ear of a healthy volunteer with our custom, handheld ME-OCT system.B, temporal bone CT (coronal view) of a patient’s healthy left ear. C, OCT 3D *en face* view of a healthy middle ear (ME) with the tympanic membrane (TM) digitally removed. A dashed orange circle shows the field of view (FOV) of the ME-OCT image. D, OCT coronal view (slice A=18.0388mm in supplementary materials). E, CT coronal view (slice A=18.0388mm). F, fused coronal OCT/CT image with CT in greyscale and OCT in false colour overlay (slice A=18.0388mm). Abbreviation: CP, cochlear promontory; H, hypotympanum, LPI, long process of the incus; M, malleus; OW, oval window; PS, Prussak’s space; RW, round window; Sc, scutum; ST, stapedius tendon; TM, tympanic membrane; U, umbo. Anatomical directions are denoted as left(L)/right(R), superior(S)/inferior(I), anterior(A)/posterior(P). Note, panel A has been intentionally left blank in accordance with medRxiv publication requirements that people or of identifying body parts should be removedReaders who wish to access the complete version of this figure are encouraged to visit the homepage of the Interactive ME-OCT Atlas at [earlab.ca]^29^ or to contact the corresponding author for further assistance.

The field of view (FOV) of ME-OCT is limited by the scanning geometry and ear canal anatomy^6,7,9^. In particular, the location and prominence of the scutum (Sc) usually obscures the epitympanum and the heads of the incus (I) and malleus (M), and in some individuals may extend far enough to significantly obscure the long process of the incus(I) and/or the manubrium of the malleus (Figure 1B). A typical 3D volumetric image of a healthy middle ear seen along the line of sight down the ear canal with ME-OCT is shown in Figure 1C. The TM has been digitally removed to provide an unobstructed view into the middle ear. A representative middle ear OCT slice from a volumetric image and the corresponding CT slice at the same orientation are shown in Figure 1D and Figure 1E. A fused image showing the CT image as a greyscale background and the OCT image as a false colour overlay is shown in Figure 1F. This presentation of fused images will be used throughout the study.

## Methods

### Case Selection and High-Resolution CT Acquisition

The study was performed at a hospital tertiary otology clinic under approval from the hospital’s Research Ethics Board (File #1019922). Informed consent was obtained in accordance with the Helsinki Declaration (JAMA 2000; 284:3043-3049). Three participants, two male (56-60 years and 51-55 years), one female (26-30 years), who had undergone high-resolution temporal bone CT scans as part of their standard care were included. High-resolution CT images without contrast were acquired by scanning the head axially from the skull vertex to the base. Details of the CT scanning configurations are summarized in Table 1.

**Table 1.** Demographic Information and CT Imaging Configuration Used for Each Patient.

| Demographic Information |  | CT Imaging Protocols |  |  |  |  |  |
| --- | --- | --- | --- | --- | --- | --- | --- |
| Case Description | Age, Y/Sex | CT Model | Voltage, Kvp | Current Rating, ma | Collimation Width, mm | Beam Pitch | Slice Thickness, mm |
| Normal Ear | 56-60 /M | Siemens Somatom Definition As+ | 120 | 163 | 0.3 | 0.85 | 0.4 |
| Traumatic Injury | 26-30 /F | Siemens Somatom Definition Flash | 120 | 164 | 0.6 | 0.85 | 0.6 |
| Chronic Cholesteatoma | 51-55 /M | Siemens Somatom Definition Edge | 120 | 127 | 0.3 | 0.85 | 1.0 |

### ME-OCT Acquisition

Prior to OCT measurement, patients had their ear canals cleared of cerumen and other debris. With the patient and operator both sitting, as shown in **Figure 1**A, the operator inserted the OCT handpiece speculum into the ear canal, centered the image on the TM under video otoscopic guidance and pressed a foot pedal button to initiate the collection of volumetric data^29,30^. During acquisition, the OCT beam was scanned over the eardrum surface in a spiral to collect 262,000 image lines distributed roughly uniformly over the TM in 4.5 seconds^9,29,30^. Data was processed offline using a custom denoising algorithm^18^ to remove artifacts and the image was exported to DICOM format.

### Image Fusion and Evaluation

The OCT and CT DICOM datasets were imported into 3D Slicer^31^ for co-registration and fusion. CT images were cropped, rescaled^32,33^ and mapped to a −1000 to 2000 Hounsfield Units (HU) greyscale range. OCT amplitude data was log-compressed and mapped to a 0-255 greyscale range. A semi-automated, intrinsic, rigid registration was performed using the SlicerIGT^34^ fiducial registration module. Bony landmarks (e.g. scutum (Sc), umbo (U), annulus, lenticular process of the incus (I) and prominences on the cochlear promontory (CP)) which were visible in both CT and OCT images were manually identified. Fused images were generated by overlaying a semi-transparent false colour OCT image onto the corresponding greyscale CT image (Figure 1F).

### Co-Registration Accuracy and Reproducibility

For each ear, we independently co-registered the CT and OCT datasets six times. We estimated the registration accuracy using the root mean square error (RMSE)^35^ between the selected landmark pairs used in the registration. To measure the reproducibility of the co-registration process we^19^:

1. Co-registered each of the three OCT volumes onto the corresponding CT volume.
2. Performed scalar volume addition of OCT and CT volumes.
3. Applied threshold-based segmentation to the fused OCT/CT volume.
4. Identified the boundary pixels of each segmented volume.
5. Selected all pairs of CT volumes (15 unique pairs between the 6 volumes per dataset × 3 datasets = 45 pairs) and calculated the maximum, 95% and mean Hausdorff distances^33,36^ between boundary pixels in the two volumes.
6. Calculated the Dice similarity coefficient (DSC)^32,36,37^ between each pair of volumes including all pixels within each volume.

An interactive website^29^, along with 3D Slicer scene files containing co-registered CT and OCT image datasets, are available as supplementary materials accompanying this study^38^. In the captions for **Figure 2, Figure 3**, and **Figure 4** coronal slices are labelled with the location along the anterior/posterior (A/P) axis, sagittal slices with the location along the patient right/left (R/L) axis and axial slices with their location along the superior/inferior (S/I) axis.

**Figure 2.**
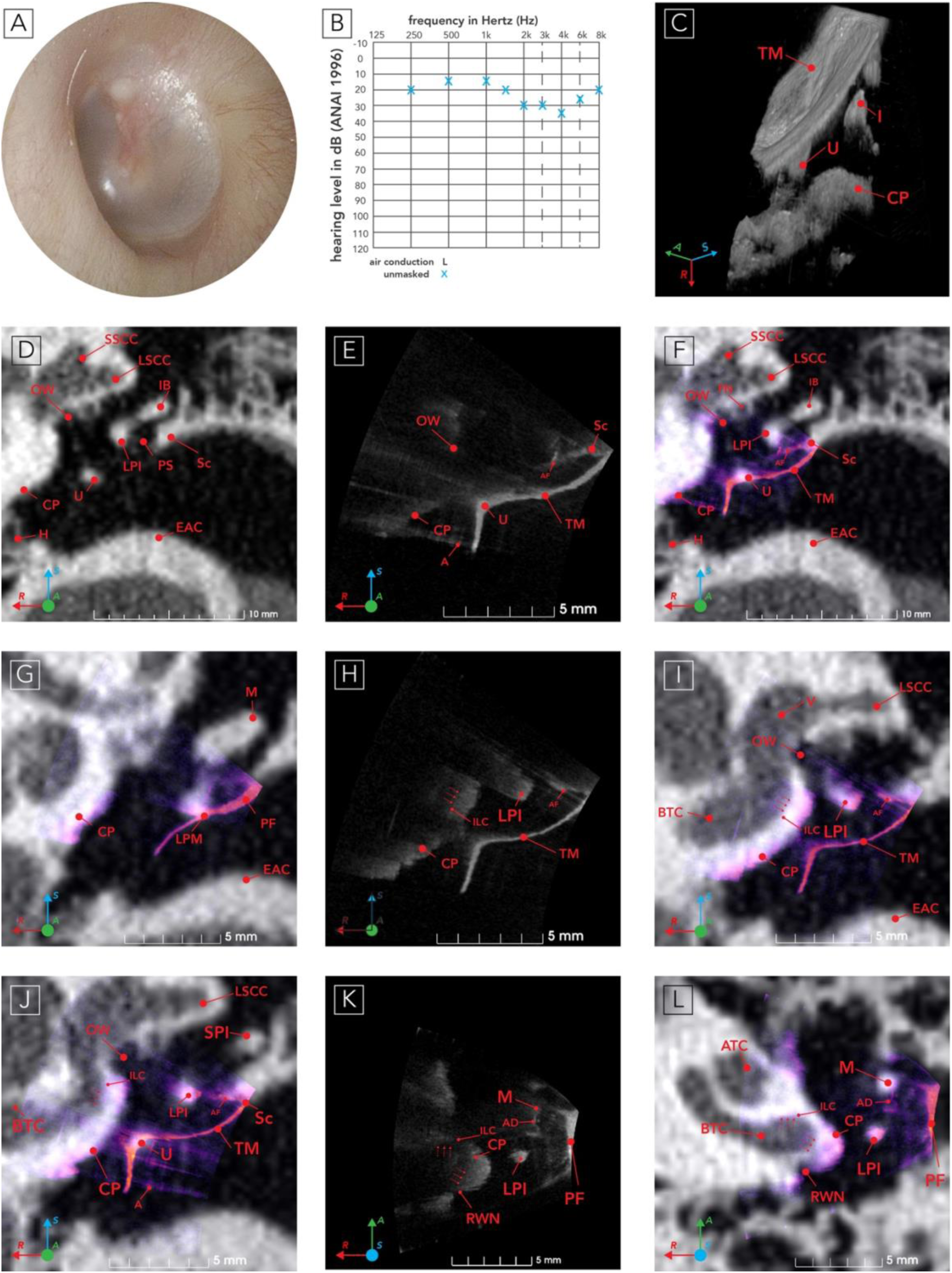
Clinical CT, OCT and OCT/CT Fusion of a Normal Ear. A, otoscopic view of the tympanic membrane obtained with a Welch-Allyn Macroview® otoscope. B, audiogram. C, 3D transtympanic OCT image. D, CT coronal view (A=18.9045mm). E, OCT coronal view (A=18.9045mm). F, fused OCT/CT coronal view (A=18.9045mm). G, fused OCT/CT coronal view (A=20.0666 mm). H, OCT coronal view (A=17.4167 mm). I, fused OCT/CT coronal view (A=17.4167 mm); (J) fused OCT/CT coronal view (A=18.2404mm). K, OCT axial view (S=8.8751 mm). L, fused OCT/CT axial view (S=8.8751 mm). Abbreviation: A, artefact; AD, adhesion; AF, accessory fold; ATC, apical turn of cochlea; BTC, basal turn of cochlea; CP, cochlear promontory; EAC, external auditory canal; H, hypotympanum; IB, incus body; I, incus; ILC, interior lumen of the cochlear duct; LPI, long process of incus; LPM, lateral process of malleus; LSCC, lateral semicircular canal; M, malleus; OW, oval window; PF, pars flaccida; PS, Prussak space; RWN, round window niche; Sc, scutum; SPI, short process of incus; SSCC, superior semicircular canal; TM, tympanic membrane; U, umbo; V, vestibule. Three red arrows highlight the interior wall of the cochlear lumen.

**Figure 3.**
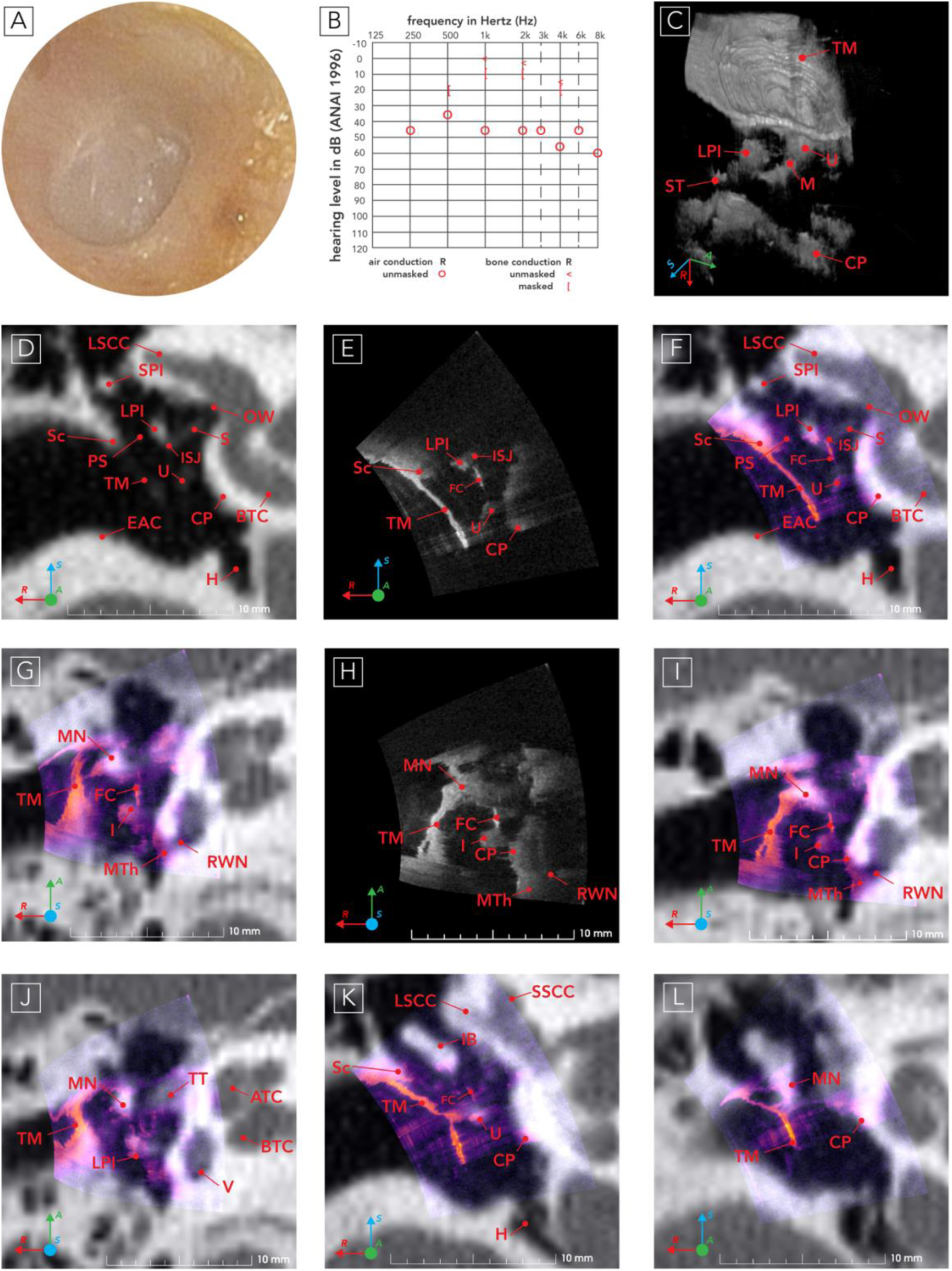
Clinical Temporal Bone CT and Middle Ear Fusion of a Traumatic Right Ear. A, endoscopic view captured with an Olympus Exera III surgical endoscopy system. B, audiogram. C, 3D transtympanic OCT. D, CT coronal view (A=186.1830mm). E, OCT coronal view (A=186.1830mm). F, fused OCT/CT coronal view (A=186.1830mm). G, fused OCT/CT axial view (S=96.8115mm). H, OCT axial view (S=95.2822mm). I, fused OCT/CT axial view (S=95.2822mm). J, fused OCT/CT axial view (S=97.3213mm). K, fused OCT/CT coronal view (A=187.3927mm). L, fused OCT/CT coronal view (A=189.4116mm). Abbreviation: ATC, apical turn of cochlea; BTC, basal turn of cochlea; CP, cochlear promontory; EAC, external auditory canal; FC, fibrous connection; H, hypotympanum; I, incus; IB, incus body; ISJ, incudostapedial joint; LPI, long process of incus; LSCC, lateral semicircular canal; M, malleus; MN, malleus neck; MTh, thickened mucosa; OW, oval window; PS, Prussak’s space; RWN, round window niche; S: stapes; Sc, scutum; SPI, short process of incus; SSCC, superior semicircular canal; ST, stapedius tendon; TM, tympanic membrane; U, umbo; V, vestibule.

**Figure 4.**
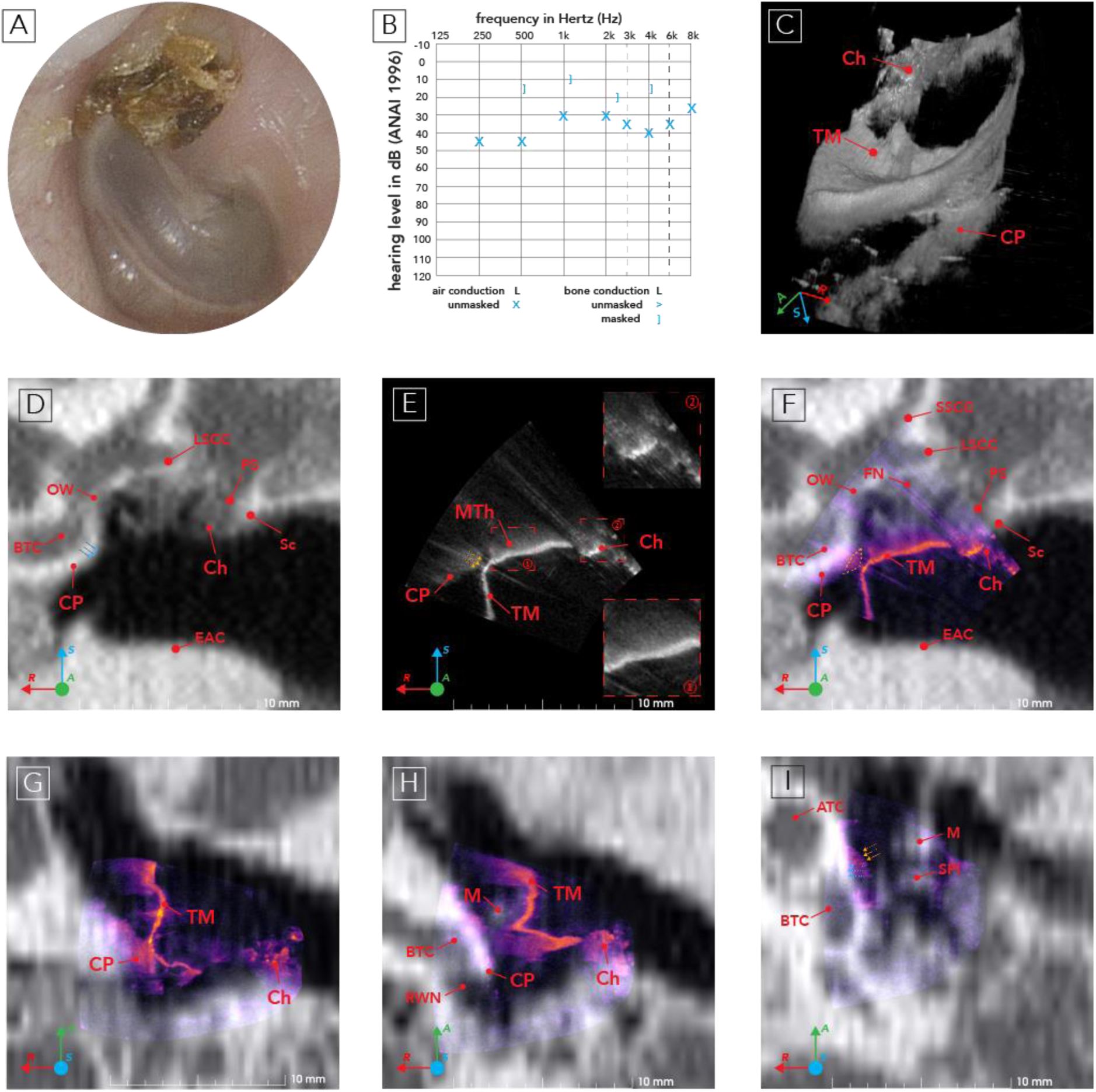
Clinical Temporal Bone CT and Middle Ear Fusion of a Left Ear with Cholesteatoma. A, Endoscopic tympanic membrane image captured with an Olympus Exera III surgical endoscopy system. B, Patient audiogram showing mild conductive loss in the left ear. C, 3D transtympanic OCT. The large black region on the TM is caused by shadowing by the cholesteatoma proximal to the image frame. D, CT coronal view (A=204.5090mm); E, OCT coronal view (A=204.5090mm) with yellow arrows highlighting the extent of cholesteatoma; F, fused OCT/CT coronal view (A= 204.5090mm), the yellow dashed triangle highlights where soft tissues that can be seen from OCT are absent from the CT image owing to a six month delay between when the CT and OCT images were acquired. G, fused OCT/CT axial view (S=1402.2731mm). H, fused OCT/CT axial view (S=1403.5721mm). I, fused OCT/CT axial view (S=1405.2535mm) with yellow arrows highlighting the extent of cholesteatoma and blue arrows highlighting the contour of the promontory. Abbreviations: ATC, apical turn of cochlea; BTC, basal turn of cochlea; Ch, cholesteatoma; CP, cochlear promontory; EAC, external auditory canal; FN, facial nerve; LSCC, lateral semicircular canal; M, malleus; MTh, thickened mucosal; OW, oval window; PS, Prussak’s space; RWN, round window niche; Sc, scutum; SSCC, superior semicircular canal; TM, tympanic membrane.

## Results

The average RMSE between landmarks in the CT and OCT datasets was 0.100 ± 0.035. Threshold-based segmentation of the fused images showed a maximum Hausdorff distance across boundary pixels in the six co-registrations of 0.95 ± 0.23 mm, a mean Hausdorff distance of 0.045 ± 0.011 mm and a 95% Hausdorff distance of 0.17 ± 0.02 mm. The mean Dice Similarity Coefficient (DSC) between the CT and OCT segmented volumes was 0.747 ± 0.061. These results indicate that this method can reliably co-register volumetric middle ear OCT images to temporal bone CT images^39,40^.

### Case I: Normal Ear

A male in the 56–60-year age range, with a mild sensorineural loss (Figure 2B) but normal left middle ear was selected as a healthy control for OCT/CT image fusion. Otoscopic examination of the left TM (Figure 2A) showed it to be intact and healthy. In the otoscopic image, the malleus (M) is visible from the umbo (U) to the short process (SPI), and the incudostapedial joint (ISJ) can be seen posteriorly through the TM (Figure 2A). The coronal CT image (Figure 2D) shows a broad external auditory canal (EAC) and an aerated middle ear space with intact scutum (Sc) and ossicles. The TM is not visible in the CT image due to its thinness and low radiopacity. The corresponding OCT image, shown alone in Figure 2E and superimposed on the co-registered CT in Figure 2F clearly shows the TM and proximal surfaces of the cochlear promontory (CP) and incus (I). Other identifiable soft tissue structures include an accessory fold (AF)^2^ (Figure 2E, Figure 2F, Figure 2H, Figure 2I, Figure 2J) between the malleus (M) and the scutum (Sc) and an adhesion (AD) attached to the anterior malleus (M) (Figure 2K, Figure 2L). As compared to the CT image, the prominence of these soft tissue structures in the OCT image is striking. Also noteworthy is the inability of OCT to resolve the distal edge of the incus (I) due to the limited penetration of OCT into bone, the shadowing of the portion of the cochlear promontory (CP) lying distal to the umbo (U) and image artefacts (A) running axially from bright features on the TM^33,41^ (Figure 2J). Feature edges appear sharper in the OCT image than in the CT image owing to the higher resolution of OCT (∼50μm) as compared to CT (∼250μm)^7^. The limited FOV of the OCT image can be seen in the abrupt termination of the inferior aspect of the TM and the inability to observe structures superior and medial to the scutum (Sc) in the epitympanum in Figure 2E and Figure 2F. Other views of the ear are presented in Figure 2G through Figure 2L. Notable features visible in the OCT images include the oval window (OW) visible in Figure 2F, Figure 2I, Figure 2J, and the round window niche (RWN) visible in Figure 2K and Figure 2L. The OCT image shows a clear difference in thickness between the pars flaccida (PF) (Figure 2G) and the pars tensa (Figure 2F, Figure 2H, Figure 2J)^42^. Although OCT’s ability to penetrate into bone is limited, parts of the otic capsule are thin enough that the OCT image traces the outline of the interior lumen of the cochlea (ILC) in Figure 2I to Figure 2L.

### Case II: Traumatic middle ear injury I

Figure 3 shows the right ear of a female participant (26–30-year age range), which was subject to a serious traumatic ear injury. The participant was urgently referred to our clinic in their late teens due to bloody ear discharge following temporomandibular joint arthroscopy. Upon examination, an anterior canal wall laceration and traumatic TM perforation were identified. Audiometric testing showed moderate conductive hearing loss, with a mean air bone gap for the frequencies 0.5-4kHz of 32.5dB (Figure 3B). Although the perforation healed, the conductive hearing loss (CHL) persisted. The patient opted not to have surgery on the ear, but has been followed by our clinic, and so her ear remains in the state it was in following her initial injury.

Examination of the patient’s healed TM shows it to be nearly featureless with the lateral process of the malleus (M) being the sole notable landmark visible under endoscopy (Figure 3A). 3D OCT imaging was performed (Figure 3D). In the CT, OCT and fused coronal images (Figure 3D, Figure 3E, and Figure 3F), it can be seen that the umbo (U) had detached from the TM and the malleus (M) had shifted medially by approximately 1.5mm. The umbo(U) of the malleus(M), which would be contiguous with the TM in a normal ear, can be seen lying inferior to the incudostapedial joint (ISJ) in the coronal cut (Figure 3F) and can be followed superiorly (Figure 3G). The umbo (U) can also be seen in axial planes (Figure 3I).

In the OCT image of Figure 3J, but not in the co-registered CT image, a fibrous connection (FC) is visible between the medialized neck of the malleus (NM) and the incus (I). Both the CT and OCT images (Figure 3J) also show a thickening of the mucosa (MTh) around the RWN and on the promontory (CP), presumably due to scarring associated with reactive inflammatory changes from persistent exposure of the mucosa (MTh) to air immediately following the injury.

The medialization of the malleus (M) was not noted in the patient’s radiology report of high-resolution temporal bone CT performed after the injury which described the ear as “unremarkable”. Presumably the lack of a clearly visible TM in the CT image made it difficult for the radiologist to notice the abnormal location of the manubrium. Because the TM is so prominent on the OCT image, the medialization of the manubrium and its disconnection from the TM is obvious in both the OCT-only image (Figure 3E) and in the fused OCT/CT images of Figure 3F, Figure 3G, and Figure 3IFigure 3. This points to a possible role for fused OCT/CT images in traumatized ears in improving the visibility of soft tissue landmarks to aid in the interpretation of the CT images given that normal anatomical relationships used to identify structures may not hold.

### Case III: Cholesteatoma (L)

A male patient in the 51–55-year age range with a history of cholesteatoma (Ch) surgery in his right ear was brought in for follow-up imaging and a hyperintensity was noted in the diffusion-weighted magnetic resonance image (MRI) in his left, unoperated ear. Upon endoscopic examination of the left ear (Figure 4A), the TM displayed a crust with flares of keratin debris at the pars flaccida (PF), revealing an attic cholesteatoma (Ch). In the endoscopic image, the manubrium and umbo are clearly visible, and the posterosuperior aspect of the TM is bulging out. The patient has also developed a mild conductive loss in his left ear as shown in the audiogram (Figure 4B).

The OCT, CT and fused coronal views (Figure 4D, Figure 4E, and Figure 4F) along with the axial images (Figure 4I to Figure 4K) all show clear evidence of TM thickening and retraction and of the contact between the TM and the inflamed mucosa (MTh) over the promontory (CP) in Figure 4J. The keratinous outer layers of the cholesteatoma (Ch) visible from the ear canal create a hyper-reflective layer overlaying a less reflective inner layer under OCT^10,44^. This hyper-reflective outer layer has previously been reported as a distinctive feature of cholesteatoma (Ch) in OCT imaging that can be used to differentiate cholesteatoma (Ch) from inflamed mucosa (MTh) ^44^. Comparison between the OCT and CT images (Figure 4E and Figure 4F) show that the OCT image delineates the inferior edge of the cholesteatoma sac in the mesotympanum. The OCT image shows a region of inflamed mucosa (MTh) superior to the TM in (Figure 4F, Figure 4I and Figure 4K) with a uniform appearance distinct (Figure 4E, inset 1) from the more irregular intensity distribution of the cholesteatoma (Figure 4E, inset 2). The most inferior portion of the TM (Figure 4F) is of normal thickness with no evidence of inflammation. Figure 4G shows that at its most medial point the TM has retracted sufficiently that it contacts the promontory mucosa. This is not evident from the corresponding CT image.

## Discussion

The three ears imaged in this study show the complementarity of OCT and CT middle ear imaging. The TM, which is often not visible above noise floor in CT images is well resolved in OCT. Other thin, soft tissue structures readily visible in OCT images but difficult to visualize in CT include adhesions (Ad), fibrous connections (FC), mucosal folds and membranes. OCT also provides differentiation of cholesteatoma (Ch) and inflamed mucosa (MTh), each of which has a distinct appearance. Though not present in the images of this study, middle ear effusion has been shown to also have a distinct appearance in OCT images quite different from cholesteatoma and mucosa^43^. Effusion, cholesteatoma and mucosa are very difficult to distinguish in CT.

OCT’s limited field of view (FOV), susceptibility to obstruction and limited penetration in tissue means that it cannot replace CT imaging for many applications. However, this study highlights that OCT can visualize many of the bony and soft tissue structures that are important in middle ear diagnostics, and because OCT does not expose patients to ionizing radiation it may be a preferable imaging modality in situations where radiation risk is of particular concern, such as in screening^26,44^, primary diagnostics^22,45^, longitudinal monitoring^23^ and paediatrics^45,46^. Co-registered OCT and CT imaging could enhance intraoperative localization of anatomical landmarks, potentially benefiting robotic surgical applications in otology^47–49^.

Though not used in this study, ME-OCT system can also be readily integrated with OCT Doppler vibrometry which offers capabilities similar to laser Doppler vibrometry for obtaining additional, complementary functional diagnostic information co-registered with OCT imaging^16,21,23,35,37,41,50^.

ME-OCT remains in its infancy and image quality is likely to improve as the technology matures. For example, the FOV of OCT could likely be extended into the epitympanum with an angled probe like those used in endoscopic ear surgery^6^. ME-OCT is also likely to benefit from advancements in contrast enhancement^51^, artefact reduction^18^ and machine learning-based image enhancement^52^ currently under development.

### Conclusion

This study presents clinical middle ear OCT images in the context of co-registered CT images in normal and pathological ears for the first time. The CT and OCT co-registration shows how the excellent soft tissue visualization of OCT can complement CT’s unobstructed, volumetric view of the middle ear and shows clearly which features of the middle ear can and cannot be visualized with OCT. The fused images may offer new insight into middle ear disorders and are useful in interpreting OCT images by putting them into the familiar context of CT. OCT’s ability to visualize and differentiate soft tissue, its lack of ionizing radiation, its real-time framerate and its relatively low cost make it a promising technology for clinical middle ear diagnostics.

## Data Availability

All unlabelled and labelled CT and OCT composite images are publicly available from an Interactive ME-OCT Altas [earlab.ca]. All data and materials, including 3D Slicer scene files for this study, are available from FigShare[Wang J, COUVREUR F, Ghedia R, et al. Scene files for ME OCT/CT fusion of three study participants. Published online 2024:939558784 Bytes. doi:10.6084/M9.FIGSHARE.26866924.V3].

https://earlab.ca/

https://figshare.com/articles/dataset/Scene_files_for_OCT_CT_fusion_of_three_study_participants/26866924

https://figshare.com/articles/media/Interactive_ME-OCT_Altas/26867023?file=48869593

## Author Contributions

Mr. Wang and Dr. Couvreur contributed equally as co-first authors. Mr. Wang and Dr. Adamson had full access to all the data in the study and take responsibility for the integrity of the data and the accuracy of the data analysis.

Concept and design: Wang, Adamson.

Acquisition, analysis, or interpretation of data: Wang, Couvreur, Farrell, Ghedia, Morris, Adamson.

Drafting of the manuscript: Wang, Couvreur, Ghedia, Morris, Adamson.

Critical revision of the manuscript for important intellectual content: All authors.

Administrative, technical, or material support: Farrell, Morris, Adamson. Supervision: Morris, Adamson.

## Credits

We acknowledge the help we received from Ms. Mary McSweeney, Department of Diagnostic Radiology, Dalhousie University, in accessing CT datasets. We also acknowledge Ms. Carla Roberts, Division of Otolaryngology–Head & Neck Surgery, Dalhousie University, for her help in patient recruitment for the OCT imaging study.

## Data Sharing Statement

All unlabelled and labelled CT and OCT composite images are publicly available from an Interactive ME-OCT Altas [earlab.ca]^29^. All data and materials, including 3D Slicer scene files for this study, are available from FigShare^38^.

